# The CRP-to-Uric Acid Index (CURI): A Novel Inflammatory–Metabolic Index to Enhance Noninvasive Screening for MASLD and Liver Fibrosis

**DOI:** 10.1101/2025.09.01.25334851

**Authors:** Tien Manh Huynh, Chien Van Nguyen, Dinh Thien Ly, Hung Quoc Ha, Ong Thinh, Tran Luong Thi Vo, Thong Duy Vo, Duc Trong Quach

## Abstract

**Background:** Metabolic dysfunction-associated steatotic liver disease (MASLD) and significant liver fibrosis (SLF) require noninvasive screening. This study evaluates the C-reactive protein-to-uric acid index (CURI), combining inflammation and metabolic dysfunction, as a biomarker for early detection of MASLD and SLF.

**Methods:** Using NHANES 2017–2020 data (n=6,687), participants were divided into training and validation cohorts. CURI was calculated as high-sensitivity C-reactive protein (hs-CRP; mg/L) × [uric acid (mg/dL)]^3^. Associations with MASLD and SLF were analyzed via logistic regression, restricted cubic splines, and mediation analysis considering insulin resistance and adipose indices. Diagnostic performance was evaluated using the area under the curve (AUC), decision curve analysis (DCA), comparing CURI with established indices: fatty liver index (FLI), hepatic steatosis index (HSI), and fibrosis-4 (FIB-4).

**Results:** MASLD and SLF prevalence were 49.6% and 10.7%, respectively. CURI was independently associated with MASLD and SLF. Nonlinear relationships were observed. Combining CURI with FLI, HSI, or FIB-4 improved diagnostic performance compared to using them alone (AUC for MASLD: 0.852 with FLI, 0.819 with HSI, 0.690 with FIB-4; AUC for SLF: 0.811 with FLI, 0.750 with HSI, 0.699 with FIB-4, p<0.001). DCA showed net benefit for MASLD. Mediation analysis show 98.1% (MASLD) and 84.8% (SLF) of effects.

**Conclusion:** CURI is a promising, cost-effective biomarker for MASLD/SLF risk stratification.

**Highlights:** *WHAT IS KNOWN:*

- MASLD and SLF require noninvasive screening.
- Existing methods like liver biopsy are invasive.
- Indices like FLI, HSI, and FIB-4 are commonly used for screening.

*WHAT IS NEW HERE:*

- CURI is a novel biomarker combining inflammation and metabolic dysfunction for screening.
- CURI is independently associated with MASLD and SLF.
- CURI enhances prediction when combined with FLI or HSI.
- Insulin resistance mediates most of CURI effect on MASLD and SLF.
- CURI is a cost-effective tool for risk stratification in resource-limited settings.

## Introduction

Metabolic dysfunction-associated steatotic liver disease (MASLD) affects approximately 1 billion individuals worldwide, paralleling the global rise in metabolic disorders such as obesity and type 2 diabetes.^1,2^ MASLD emphasizes the critical role of metabolic factors and refines the impact of factors such as viral infections and alcohol on diagnosis.^3^ This chronic liver condition, driven by metabolic dysfunction, can progress from simple steatosis to severe outcomes, including significant liver fibrosis (SLF), cirrhosis, and hepatocellular carcinoma.^4^ These advanced stages significantly increase mortality and impose substantial health and economic burdens.^3^ The current gold standard for diagnosing MASLD and assessing fibrosis, liver biopsy, is limited by its invasiveness, high cost, sampling errors, and poor patient acceptability.^4^ Consequently, there is an urgent need for noninvasive, cost-effective biomarkers to detect MASLD and monitor its progression. Recent evidence has elucidated the underlying mechanisms of insuline resistance and inflammation in the onset and progression of MASLD.^4,5^ Central to the pathogenesis of MASLD is inflammation, which can trigger oxidative stress, lipid peroxidation and the activation of immune cells.^3^ According to recent studies, serum uric acid (SUA) is the final metabolite of purines, and elevated SUA levels can contribute to insulin resistance through various mechanisms, ultimately leading to metabolic syndrome and MASLD.^6-8^ Individuals with NASH have increased levels of high-sensitivity C-reactive protein (hs-CRP) and inflammatory cytokines, which may contribute to chronic inflammation and facilitate disease progression.^9,10^ Noninvasive indices like FLI, HSI, and FIB-4 are widely used for detecting steatotic liver disease and fibrosis, with FLI showing strong accuracy for fatty liver, HSI and FLI effective for MASLD detection, and FIB-4 remaining a validated tool for estimating fibrosis, especially in primary care.^11-14^ However, these indices are predominantly based on metabolic and anthropometric parameters and may lack sensitivity in capturing systemic inflammation or insulin resistance—key mechanisms in MASLD progression.^14^ To date, no study has assessed the predictive value of the CURI, which combines inflammatory and metabolic signals. Our study evaluated whether CURI can enhance risk stratification—alone or in combination with established scores—and thus serve as a pragmatic, accessible biomarker for MASLD and SLF, especially in resource-limited settings.

## Materials and methods

### Study design and participants

This cross-sectional study used data from the National Health and Nutrition Examination Survey (NHANES) cycles from 2017--March 2020, a nationally representative survey conducted by the National Center for Health Statistics (NCHS) under the Centers for Disease Control and Prevention (CDC). The NHANES employs a multistage, stratified probability-cluster sampling design to assess the health and nutritional status of the US civilian noninstitutionalized population. The survey combines interviews, physical examinations, and laboratory tests, with approximately 5000 participants annually. All the data are publicly available.^15,16^ We included adults aged ≥18 years who underwent vibration-controlled transient elastography (VCTE) during the specified cycles. The exclusion criteria for pregnant women were excessive alcohol consumption (>14 standard drinks/week for women or >21 for men), incomplete or missing VCTE data (including median liver stiffness measurement [LSM] or controlled attenuation parameter [CAP]), autoimmune hepatitis, missing covariate data, or insufficient data for calculating hs-CRP and SUA levels. The study was approved by the NCHS Ethics Review Board, and all participants provided written informed consent.

### Bimarker measurement

hs-CRP levels were quantified via a particle-enhanced turbidimetric immunoassay. SUA concentrations were determined via a specific endpoint enzymatic reaction, with the resultant product measured photometrically at a wavelength of 546 nm. For this study, the CRP-Uric Index (CURI) was calculated as hs-CRP × uric acid, where hs-CRP was expressed in mg/L and uric acid in mg/dL. The value of uric acid was first divided by 1000 to ensure consistency in units before multiplication.

Within the study’s methodological framework, we expressed CURI as hs-CRP (mg/L) × [SUA (mg/dL)]^3^, which was identified through a systematic feature engineering and screening process.^17^ CURI was selected through systematic feature engineering, in which 1920 potential predictors derived from mathematical transformations (including identity and cubic forms) and pairwise combinations of hs-CRP and uric acid were evaluated. Selection prioritized the highest area under the receiver operating characteristic curve (AUC) in univariate logistic regression, favouring interpretability and discriminative power. For regression models, CURI was rescaled by dividing by 100 to improve numerical stability. All ROC cut-offs and thresholds are reported on the unscaled metric unless otherwise stated.

For comparison, established indices were calculated as follows: HSI = 8 × (alanine aminotransferase [ALT]/aspartate aminotransferase [AST] ratio) + body mass index (BMI) (+2 if female; +2 if diabetes mellitus present), with HSI >36 indicating steatosis; and FIB-4 = (age [years] *AST)/(platelet count [10^9^/L] * ALT^(0.5)^).^12,14,18^

### Assessment of hepatic steatosis and fibrosis

Hepatic steatosis and fibrosis were evaluated via VCTE with a FibroScan® model 502 V2 Touch (FibroScan, Echosens, Paris) equipped with medium or extralarge probes. Trained technicians performed examinations, which required ≥10 valid measurements with an interquartile range (IQR)/median ratio, with higher values indicating more severe fibrosis, whereas CAP measured ultrasonic attenuation in the liver. CAP values range from 100–400 dB/m, with higher values indicating greater amounts of liver fat. SLD was characterized by a median CAP□≥□275 dB/m with elastographic evidence of steatosis, whereas CSF was identified by a median LSM□≥□8.0 kPa, as indicated by prior studies.^19-21^

MASLD was defined by the presence of liver steatosis in conjunction with metabolic dysfunction while excluding other potential causes of hepatic fat accumulation. To establish metabolic dysfunction, patients must meet at least one of the following criteria: (1) a body mass index (BMI) of 25 kg/m2 or higher or a waist circumference exceeding 94 cm in men or 80 cm in women; (2) fasting blood glucose levels of 100 mg/dL or above, hemoglobin A1c values of 5.7% or greater, a prior diagnosis of type 2 diabetes, or current diabetes treatment; blood pressure readings of 130/85 mmHg or above, or ongoing antihypertensive therapy; triglyceride levels of 150 mg/dL or greater, or being under lipid-lowering treatment; and low high-density lipoprotein cholesterol (HDL-C) levels, with thresholds set at below 40 mg/dL for men and below 50 mg/dL for women, or active treatment for dyslipidemia.^22,23^

Alcohol consumption was assessed via self-report questionnaires (ALQ111, ALQ121, ALQ130), with one standard drink equaling 14 g ethanol. Excessive intake was classified as >350 g/week (women) or >420 g/week (men).

### Covariates

Demographic data (age, sex, race/ethnicity, poverty-to-income ratio, marital status, education) were collected via computer-assisted personal interviews. BMI was calculated from measured weight and height and categorized as normal/underweight (<25 kg/m^2^), overweight (25-<30 kg/m^2^), or obese (≥30 kg/m^2^). Diabetes was defined by self-reports, medication use, HbA1c ≥6.5%, or fasting glucose ≥126 mg/dL. Hypertension was based on self-reports or antihypertensive medication. Cardiovascular disease included self-reported coronary heart disease, heart attack, congestive heart failure, or stroke. Chronic kidney disease was defined as an estimated glomerular filtration rate <60 mL/min/1.73 m^2^ (MDRD formula). Smoking status was categorized as never (<100 lifetime cigarettes), former (>100 cigarettes but not current), or current. The laboratory measurements included AST, ALT, albumin, total cholesterol, HDL-C and triglycerides^23^.

### Definition of potential mediators

Mediators represented insulin resistance and adiposity: the homeostatic model assessment for insulin resistance (HOMA-IR) was calculated as fasting plasma glucose (mmol/L) × fasting insulin (mIU/L)/22.5; the metabolic score for insulin resistance (METS-IR) was defined as ln[2 × fasting glucose (mg/dL) + triglycerides (mg/dL)] × BMI (kg/m^2^)/ln[HDL-C (mg/dL)].^24^

### Statistical analysis

Analyses were conducted via R (version 4.3.2) and EmpowerStats (version 4.2), which incorporate NHANES sample weights to account for the complex survey design. Two-sided P<0.05 indicated significance. The cohort was randomly split 70:30 into training and validation sets. Continuous variables are presented as weighted means (SDs) or medians (IQRs), and categorical variables are presented as unweighted counts (weighted percentages). Differences were tested via weighted t tests/Wilcoxon rank-sum tests for continuous variables and chi-square tests for categorical variables. Multivariate logistic regression was used to assess the associations of the CURI with MASLD/SLF in three models: unadjusted (Model 1); adjusted for demographic factors (age, sex, race/ethnicity, marital status, and education; Model 2); and further adjusted for clinical/lifestyle factors (smoking, obesity, diabetes, hypertension, cardiovascular disease, chronic kidney disease, and cancer; Model 3). Dose response was explored via generalized additive models (GAMs) and two-piecewise linear regression to identify thresholds (K), with likelihood ratio tests comparing fits. Subgroup analyses and interaction terms were used to evaluate effect modification. Trends across CURI quartiles were assessed by the Cochran□Armitage test. Predictive performance was evaluated by AUC and decision curve analysis (DCA) for net benefit. Calibration was assessed graphically. Causal mediation analysis decomposed CURI effects on MASLD/SLF via HOMA-IR and METS-IR to estimate direct/indirect effects and mediation proportions by bootstrapping (1000 iterations).

## Results

### Study population

Data from 15,550 individuals in the NHANES 2017–March 2020 cycles were screened, resulting in 6,687 eligible participants after exclusions (see Supplementary Figure 1, Supplementary Digital Content 1 for participant flow), were screened, resulting in 6687 eligible participants after applying exclusion criteria (. The cohort was randomly divided into a training set (n=4680; 70%) for model development and a validation set (n=2007; 30%) for independent assessment, with no statistically significant differences in baseline characteristics between the sets (all P>0.05). MASLD prevalence was 49.6% (n=3319) and SLF prevalence was 10.7% (n=715). In both sets, participants with MASLD had higher body mass index (BMI), waist-to-hip ratio (WHR), alanine aminotransferase (ALT), and aspartate aminotransferase (AST) levels, and greater prevalence of hypertension, type 2 diabetes, chronic kidney disease, and dyslipidaemia than those without MASLD (all p <0.001, table 1A). Similarly, participants with SLF had more adverse metabolic profiles, including higher BMI, WHR, aminotransferase levels, and rates of hypertension and diabetes (all p<0.001 except where noted; table 1B).

### CURI distribution in MASLD and SLF population

CURI values were significantly elevated in participants with MASLD (median 46.8, IQR 18.7– 118.3 vs 15.1, IQR 5.7–42.1; p<0.001) and SLF (median 68.2, IQR 28.4–179 vs 24.7, IQR 8.7–68.7; P<0.001; see Supplementary Figure 2, Supplementary Digital Content 1). The distributions were consistent across the training and validation sets.

### Associations between the CURI and MASLD/SLF

Logistic regression showed that higher CURI was significantly associated with both MASLD and SLF (Table 2). In the unadjusted model, each 100-unit increase in CURI was linked to higher odds of MASLD (OR 2.31; 95% CI, 1.98–2.73) and SLF (OR 1.30; 95% CI, 1.20– 1.42), both *P* < 0.001. These associations remained significant after adjusting for demographics (Model 2: MASLD OR 2.18; SLF OR 1.26) and full covariates (Model 3: MASLD OR 1.30; SLF OR 1.14). Quartile-based analyses confirmed a dose–response relationship. Compared with individuals in the lowest quartile of CURI (Q1), those in Q2, Q3, and Q4 had progressively higher odds of MASLD, with aORs of 1.59 (95% CI, 1.29–1.95; P < 0.001), 2.42 (95% CI, 1.97–2.98; P < 0.001), and 3.06 (95% CI, 2.45–3.84; P < 0.001), respectively (p for trend < 0.001). A similar gradient was observed for SLF, with aORs of 0.97 (95% CI, 0.65–1.45; p = 0.870) for Q2, 1.63 (95% CI, 1.13–2.38; p = 0.010) for Q3, and 2.44 (95% CI, 1.70–3.56; P < 0.001) for Q4 (p for trend < 0.001).

### Nonlinear dose□response relationships

GAM revealed nonlinear patterns: an L-shaped curve for MASLD (initial steep risk increase, followed by plateau) and an inverted U-shaped curve for SLF (risk peaking at intermediate CURI levels before declining (Figure 1). Threshold effect analysis via two-piecewise linear regression identified inflection points at CURI=145.4 for MASLD and 420.6 for SLF (Table 3). Below these points, associations were strong (MASLD: aOR 2.16 per 100 units, 95% CI 1.84--2.55; P<0.001; SLF: aOR 1.31 per 100 units, 95% CI 1.20-1.43; P<0.001). Above the thresholds, the slopes attenuated (MASLD: aOR 1.00 per 100 units, 95% CI 0.97–1.03; p<0.001; SLF: aOR 1.01 per 100 units, 95% CI 0.98–1.03; p<0.001). Log-likelihood ratio tests confirmed the superior fit of the segmented models (both p<0.001) (see Supplementary table 1, Supplementary Digital Content 1).

**Figure 1:**
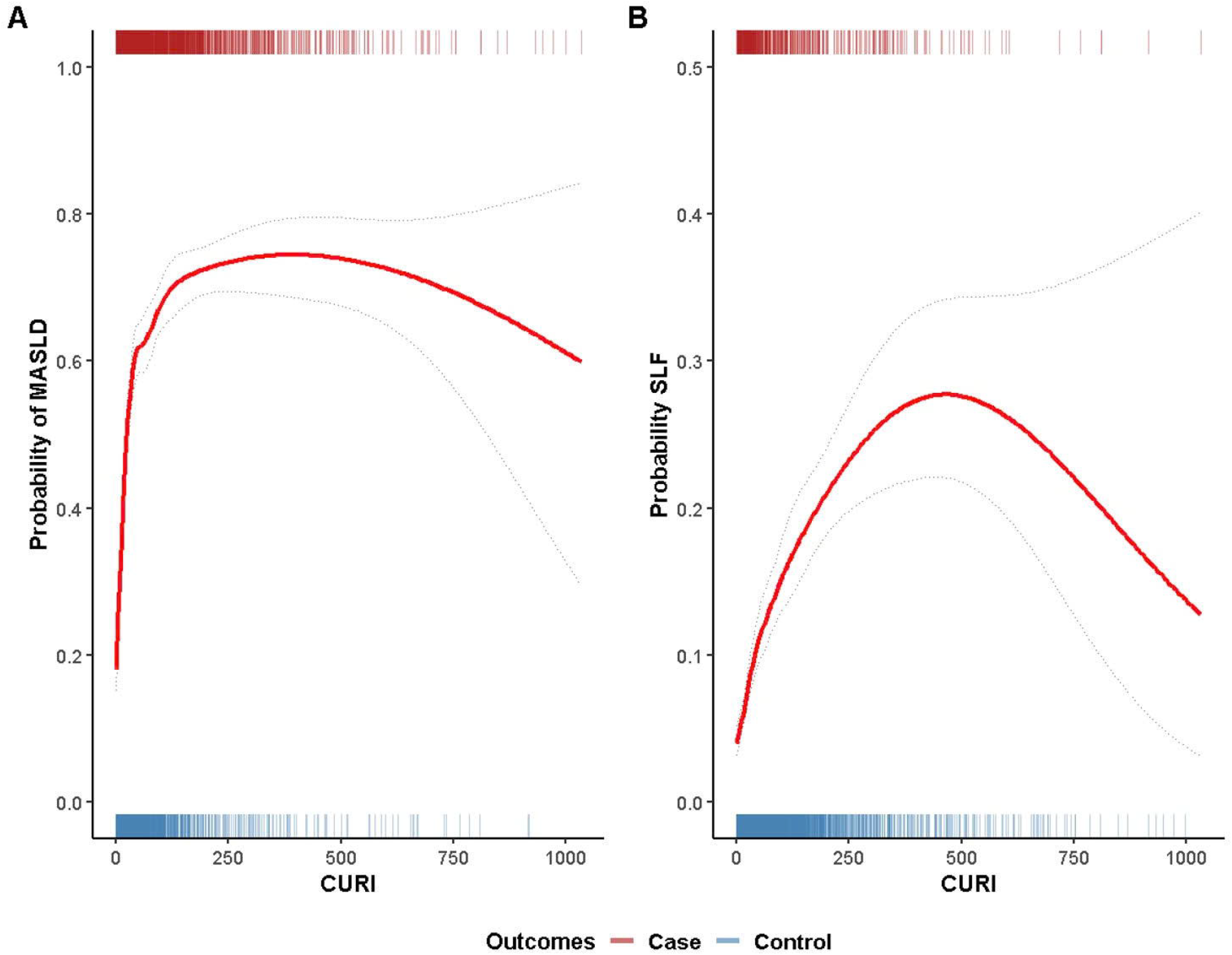
Associations of the hs-CRP/HDL-C index with MASLD (A) and SLF (B). The red solid line illustrates the smoothed curve fit between the variables, with the gray dashed lines and the area between them indicating 95% confidence intervals around the fitted values. The plot shows the adjusted probability of **(A)** metabolic dysfunction-associated steatotic liver disease (MASLD) and **(B)** significant liver fibrosis (SLF) according to the CURI. The solid red line represents the estimated probability from a restricted cubic spline model, and the dashed gray lines represent the 95% confidence intervals. In Panel **A**, the cases are participants with MASLD, whereas the controls are non-MASLD participants. Similarly, in Panel **B**, cases and controls correspond to participants with and without SLF, respectively.

### Subgroup and interaction analyses

Stratified analyses (Model 3) revealed consistent positive associations for MASLD across subgroups, with significant effect modification by age (stronger at <60 years; p interaction=0.017), sex (stronger in males; p=0.017), race/ethnicity (stronger in non-Hispanic White; P<0.001), marital status (p=0.017), education (p<0.001), smoking (p=0.005), obesity (p=0.017), hypertension (p<0.001), and chronic kidney disease (p=0.017; Figure 4). For SLF, associations varied, with significant interactions for sex (p=0.029), race/ethnicity (p=0.006), marital status (p=0.029), education (p=0.003), smoking (p=0.012), obesity (p=0.030), hypertension (p=0.004), and chronic kidney disease (p=0.030). No interactions were detected for diabetes, cardiovascular disease, or cancer (all p>0.05) (Figure 2).

**Figure 2:**
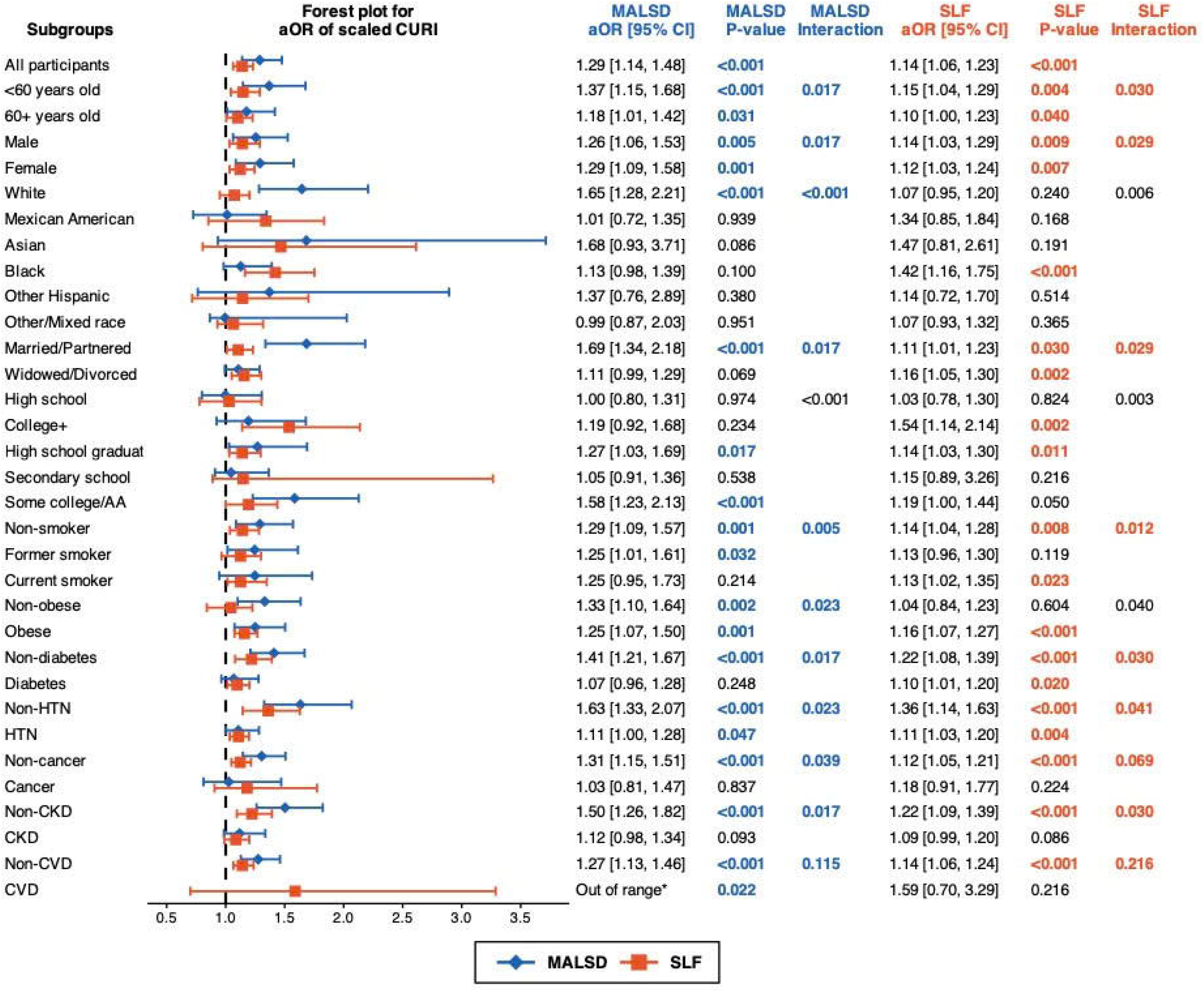
Subgroup analysis of the associations between the CURI and the odds of MASLD and SLF. The results are presented as adjusted odds ratios with 95% confidence intervals (CIs). Statistically significant associations are highlighted in bold. **Annatation**: ^*^ The extremely large aOR observed in the CVD subgroup for MASLD (aOR 44.6, 95% CI 1.20– 4.25×10^11^) likely reflects model nonconvergence due to sparse data. **Abbreviations**: aOR, adjusted odds ratio; CI, confidence interval; CKD, chronic kidney disease; CVD, cardiovascular disease; HTN, hypertension; MALSD, metabolic dysfunction-associated steatotic liver disease; SLF, significant liver fibrosis.

### Mediation analysis

Causal mediation analyses (adjusted for demographics and clinical factors) indicated that insulin resistance was a key mediator (Figure 3). For MASLD, METS-IR mediated 98.1% of the total effect (95% CI 76.1-131.2; P<0.001), with a significant indirect effect and negligible direct effect (β≈0.000). HOMA-IR was 39.2% (95% CI 10.7–83.7; P<0.001). For SLF, METS-IR mediated 84.8% (95% CI 57.9-149.9; p<0.001), and HOMA-IR mediated 7.3% (95% CI 1.7--32.7; p=0.005), again with predominant indirect effects. The supporting regressions revealed a stronger CURI association with METS-IR (model 3: β=7.1, 95% CI 5.8-8.4; p<0.001) than with HOMA-IR (β=0.40, 95% CI 0.15--0.65; P=0.002). Both mediators independently predicted outcomes, with larger effects for METS-IR (e.g., MASLD: aOR 1.12, 95% CI 1.10–1.14; p<0.001) (see Supplementary table 2, Supplementary Digital Content 1).

**Figure 3:**
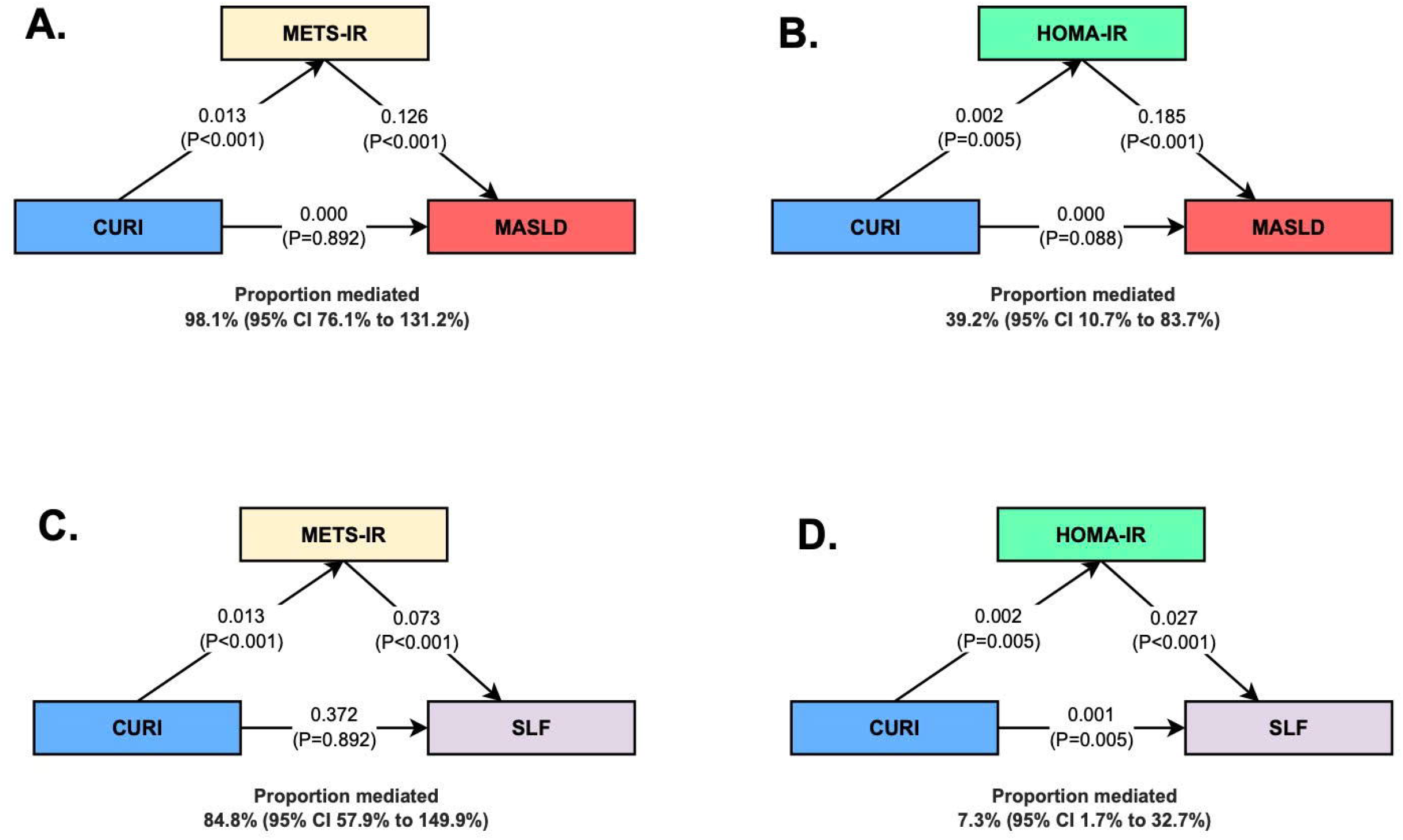
Analysis of the associations of the CURI with the MASLD score and SLF via insulin resistance markers. Path coefficients (β) from adjusted logistic regression models are shown on the arrows. A coefficient of 0.000 indicates a nonsignificant direct effect (equivalent to an odds ratio of ∼1.0). Annatation: Pathways adjusted for age, sex, race, marital status, education, smoking status, obesity, diabetes mellitus, hypertension, cancer, eGFR, chronic kidney disease, and cardiovascular disease. The proportion of the total effect mediated by HOMA-IR or METS-IR is presented with its 95% confidence interval. Abbreviations: CURI: mg/dL^4^; HOMA-IR was calculated as follows: fasting insulin (μU/mL) × fasting glucose (mmol/L)/22.5. METS-IR was calculated as (Ln[(2 × fasting glucose (mg/dL)) + fasting triglycerides (mg/dL)] × BMI)/Ln[fasting HDL-c (mg/dL)].

### Diagnostic performance

In the validation cohort, the CURI index demonstrated moderate discriminative ability for MASLD (AUC: 0.707; 95% CI: 0.685–0.730) and SLF (AUC: 0.700; 95% CI: 0.666–0.735). The optimal thresholds identified via Youden’s index were 17.385 for MASLD (sensitivity: 77.6%; specificity: 53.8%) and 51.145 for SLF (sensitivity: 63.7%; specificity: 69.0%) (Table 5). CURI significantly outperformed individual biomarkers, including hs-CRP (MASLD AUC: 0.683, *p* < 0.001; SLF AUC: 0.668, *p* = 0.002), uric acid (MASLD AUC: 0.640, *p* < 0.001; SLF AUC: 0.653, *p* = 0.003), and FIB-4 (MASLD AUC: 0.533, *p* < 0.001; SLF AUC: 0.620, *p* = 0.004). Compared with established indices, CURI yielded lower standalone performance than did HSI (MASLD AUC: 0.817, *p* < 0.001 vs CURI; SLF AUC: 0.748, *p* = 0.024 vs CURI) but demonstrated superior predictive value when integrated into composite models. The combination of the CURI with FLI produced the highest discrimination for both outcomes (MASLD AUC: 0.852, 95% CI: 0.829–0.876; SLF AUC: 0.811, 95% CI: 0.768–0.854), with both combinations significantly outperforming the CURI alone (*p* < 0.001 for both) (Table 3).

### Clinical utility and calibration

DCA demonstrated a net clinical benefit for the CURI in MASLD prediction at risk thresholds <0.35 (comparable to the HSI/FLI at higher thresholds) and a limited benefit for the SLF across narrower thresholds (see Supplementary Figure 3, Supplementary Digital Content 1). The calibration plots revealed good agreement between the predicted and observed probabilities for MASLD (in particular, 0.3--0.7 range) and acceptable, although variable, patterns for SLF (minor overestimation at low probabilities) (see Supplementary Figure 4, Supplementary Digital Content 1).

## Discussion

This first study to evaluate CURI, a composite biomarker integrating inflammatory and metabolic signals, in relation to MASLD and SLF using NHANES data, found that higher CURI values were independently associated with increased odds of both outcomes. CURI outperformed hs-CRP and SUA, and its combination with FLI, HSI, or FIB-4 improved discrimination for steatosis and fibrosis. DCA showed clinical benefit for MASLD at thresholds <0.35, and limited benefit for SLF. Subgroup analyses showed consistent associations across age, sex, race/ethnicity, and other factors (p < 0.05 for interactions). Mediation analysis indicated that insulin resistance and visceral adipose tissue, especially measured by METS-IR, mediated 98.1% of the effect on MASLD and 84.8% on SLF.

Meta-analyses have shown that hs-CRP and SUA are associated with MASLD, suggesting that the CURI is a composite of inflammatory–metabolic stress.^25^ The dose–response curve was nonlinear: L-shaped for MASLD with a threshold near 145.4 and inverted-U for SLF peaking at approximately 420.6, these patterns suggest saturation phenomena that warrant mechanistic study.

MASLD pathogenesis follows a “multiple-hit” model (lipotoxicity, insulin resistance, diet, gut microbiota, genetics).^4,26^ The levels of proinflammatory cytokines (TNF-α, IL-6, and IL-1β) and CRP are consistently greater in MASLD patients than in controls, and these levels converge on stress–inflammation pathways (JNK/AP-1 and IKK/NF-κB) and link metabolic stress to inflammation.^5,27^ Clinically, the disease progresses from steatosis to MASH, fibrosis, cirrhosis, and hepatocellular carcinoma, with excess cardiovascular events and mortality, persistent inflammation is a major driver.^26,28,29^ The inflammatory process is complex, involving hepatic lipid metabolism, gut microbiome signals, toll-like receptor 4 (TLR4) activation, and adipose tissue-liver crosstalk, forming a feed-forward loop of inflammation and fibrosis.^30^ Evidence also implicates hs-CRP is related to MASLD and mediates its ability to predict 10-year ASCVD risk (14.7%).^21^ SUA is strongly linked to MetS^24^ and predicts MetS in cohorts (notably in Chinese adults).^7^ Mechanistically, xanthine oxidoreductase activity and urate lowering affect MetS features in animal models.^31^ Hyperuricaemia increased the incidence of MASLD (HR 2.168 in the “only HUA” group). ^32^ SUA activates NLRP3/IL-1β and promotes hepatic inflammation and fibrosis; gut dysbiosis may amplify oxidative stress and lipid accumulation.^33^ Urate oxidase–knockout mice further support the involvement of the ROS/JNK/AP-1 pathway in steatosis. ^25,33^ Furthermore, elevated SUA and CRP levels have each been linked to insulin resistance and central fat accumulation, supporting a plausible inflammatory-metabolic mechanism underlying liver disease progression.^34^ Although no studies have directly examined whether CURI effect on MASLD and SLF is mediated by insulin resistance and visceral adiposity, existing evidence supports this pathway. Prior analyses using HEI as an exposure demonstrated that insulin resistance (e.g., METS-IR, HOMA-IR) and visceral fat indices (e.g., BRI, ABSI) significantly mediated associations with MASLD.^35^ METS-IR has also been validated as a reliable proxy for visceral adiposity and a predictor of type 2 diabetes.^36^

Because CURI relies on routine laboratory tests, it is simple to implement and scalable for population screening and primary-care triage. Consistent with guideline-endorsed sequential algorithms, its role is adjunctive rather than replacement.^14^ When added to established indices for steatosis (HSI/FLI) or fibrosis (FIB-4), CURI improved discrimination in our validation analyses, indicating complementary information beyond traditional metabolic and anthropometric inputs. A pragmatic two-step pathway would preselect with HSI/FLI or FIB-4 and then apply CURI to refine the pretest probability before elastography or referral— particularly where imaging access is limited. Future research will specifically investigate CURI in populations with MASLD, at-risk Metabolic Dysfunction-Associated Steatohepatitis, and advanced cirrhosis. These studies will aim to calibrate context-specific cut-offs and assess whether CURI-guided pathways can reduce unnecessary imaging while maintaining high sensitivity for detecting advanced disease.

The strengths include a large, nationally representative sample, prespecified training-validation cohorts, survey-weighted analyses; assessment of discrimination, calibration, and decision curve net benefit and mediation analyses showing that IR—particularly by METS-IR—accounts for a substantial proportion of the CURI–MASLD–SLF association. Limitations include the cross-sectional design (precluding causal inference), reliance on transient elastography rather than biopsy, potential residual confounding factors (e.g., diet, genetics), and generalizability beyond the US. Acute inflammatory states and medications that modulate CRP or SUA (e.g., corticosteroids, NSAIDs/aspirin, statins, urate-lowering therapy) may transiently alter the CURI independent of liver status; sensitivity analyses excluding such exposures, or adjustments where feasible, will be informative in future studies.

In conclusion, CURI offers a cost-effective, scalable tool for MASLD and SLF screening, particularly in resource-limited settings. Its integration with FLI or HSI could optimize risk stratification, reducing unnecessary imaging while maintaining sensitivity. Further validation across diverse populations is warranted.

## Supporting information

Supplemental Digital Content 1

## Data Availability

All data produced are available online at the CDC/NCHS website.

https://wwwn.cdc.gov/nchs/nhanes/?utm_source=chatgpt.com

## Abbreviations

MASLD: Metabolic Dysfunction-Associated Steatotic Liver Disease
SLF: Significant Liver Fibrosis
CURI: C-reactive Protein-to-Uric Acid Index)
CRP: C-reactive Protein)
hs: CRP-High-Sensitivity C-reactive Protein
SUA: Serum Uric Acid
HOMA: IR-Homeostatic Model Assessment for Insulin Resistance
METS-IR: Metabolic Score for Insulin Resistance)
AUC: Area Under the Curve
ROC: Receiver Operating Characteristic
FLI: Fatty Liver Index
HSI: Hepatic Steatosis Index
FIB-4: Fibrosis-4 Index
VCTE: Vibration-Controlled Transient Elastography
CAP: Controlled Attenuation Parameter
ALT: Alanine Aminotransferase
AST: Aspartate Aminotransferase
BMI: Body Mass Index
HDL-C: High-Density Lipoprotein Cholesterol
CI: Confidence Interval
OR: Odds Ratio
aOR: Adjusted Odds Ratio
IQR: Interquartile Range

## Acknowledgement

The authors thank the National Center for Health Statistics of the Centers for Disease Control and Prevention for sharing the National Health and Nutrition Examination Survey (NHANES) data.

