## Supplemental Digital Content 1 for "The CRP-to-Uric Acid Index (CURI): A Novel Inflammatory–Metabolic Index to Enhance Noninvasive Screening for MASLD and Liver Fibrosis"

**SUPPLEMENTARY FILES**

**Supplementary table 1. Threshold effect analyses of the effect of the CURI on MASLD and SLF using a two-piecewise linear regression model.**

|  | **MASLD** | |  | **SLF** | |
| --- | --- | --- | --- | --- | --- |
| **Objects** | **Cut-off/aOR (95% CI)** | **P value** |  | **Cut-off/aOR (95% CI)** | **P value** |
| **Turning point (K)** | 145.40 |  |  | 420.59 |  |
| **CURI < K (per 100 units)** | 2.16 (1.84, 2.55) | **<0.001** |  | 1.31 (1.20, 1.43) | **<0.001** |
| **CURI > K (per 100 units)** | 1.00 (0.97, 1.03) | 0.902 |  | 1.01 (0.98, 1.03) | 0.705 |
| **LLR test** |  | **<0.001** |  |  | **<0.001** |

***Abbreviations****: aOR, adjusted odd ratio; CURI, an index calculated as hsCRP × (uric acid)³, units: (mg/dl)^4^; LLR, logarithm likelihood ratio; MASLD, metabolic dysfunction-associated steatotic liver disease; SLF, significant liver fibrosis****. Annatation****: P<0.05 are bolded*

**Supplementary table 2. Associations between Insulin Resistance Indices and Liver Outcomes**

| **Predictor-outcome pairs** | | |  | | **Model 1** | |  | **Model 2** | |  | **Model 3** | |
| --- | --- | --- | --- | --- | --- | --- | --- | --- | --- | --- | --- | --- |
| **Predictors*** | **Outcomes** |  | | **Beta/cOR (95% CI)** | | **P value** |  | **Beta/aOR (95% CI)** | **P value** |  | **Beta/aOR (95% CI)** | **P value** |
| CURI | HOMA-IR |  | | 0.69 (0.44, 0.95) | | <0.001 |  | 0.66 (0.43, 0.89) | <0.001 |  | 0.40 (0.15, 0.65) | 0.002 |
| CURI | METS-IR |  | | 6.40 (5.50, 7.20) | | <0.001 |  | 5.80 (5.00, 6.60) | <0.001 |  | 7.1 (5.8, 8.4) | <0.001 |
| HOMA-IR | MASLD |  | | 1.06 (1.05, 1.06) | | <0.001 |  | 1.06 (1.05, 1.07) | <0.001 |  | 1.03 (1.02, 1.04) | <0.001 |
| METS-IR | MASLD |  | | 1.13 (1.12, 1.14) | | <0.001 |  | 1.14 (1.12, 1.15) | <0.001 |  | 1.12 (1.10, 1.14) | <0.001 |
| HOMA-IR | SLF |  | | 1.01 (1.01, 1.01) | | <0.001 |  | 1.01 (1.01, 1.01) | <0.001 |  | 1.00 (1.00, 1.01) | 0.020 |
| METS-IR | SLF |  | | 1.07 (1.06, 1.09) | | <0.001 |  | 1.08 (1.07, 1.09) | <0.001 |  | 1.06 (1.04, 1.08) | <0.001 |

****Abbreviations:*** *CURI, Cardiometabolic Uric Acid Index, units: (mg/dL)⁴; HOMA-IR, Homeostatic Model Assessment for Insulin Resistance, calculated as: fasting insulin (μU/mL) × fasting glucose (mmol/L)/22.5; MASLD, metabolic dysfunction–associated steatotic liver disease; METS-IR, metabolic score for insulin resistance, calculated as (Ln[(2 × fasting glucose (mg/dL)) + fasting triglycerides (mg/dL)] × BMI)/Ln[fasting HDL-c (mg/dL)]; SLF, significant liver fibrosis.* ***Annotations****:* ***Model 1*** *was unadjusted.* ***Model 2*** *was adjusted for age, sex, race, and marital status.* ***Model 3*** *was further adjusted for education, smoking status, obesity status, diabetes status, hypertension status, and history of cardiovascular, renal, and malignant diseases. Linear regression models were applied for HOMA-IR and METS-IR, whereas logistic regression models were applied for MASLD and SLF.*

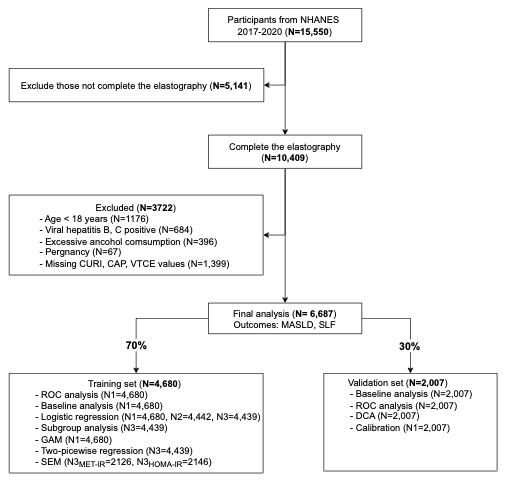

**Supplementary Figure 1. Flow chart of this study. *Abbreviation****s: NHANES, National Health and Nutrition Examination Survey; CURI, C-reactive protein-to-uric acid ratio index; CAP, controlled attenuation parameter; VCTE, vibration-controlled transient elastography; MASLD, metabolic dysfunction-associated steatotic liver disease; SLF, significant liver fibrosis; ROC, receiver operating characteristic; DCA, decision curve analysis; GAM, generalized additive model; SEM, structural equation modelling; METS-IR, metabolic score for insulin resistance; HOMA-IR, homeostatic model assessment for insulin resistance.*

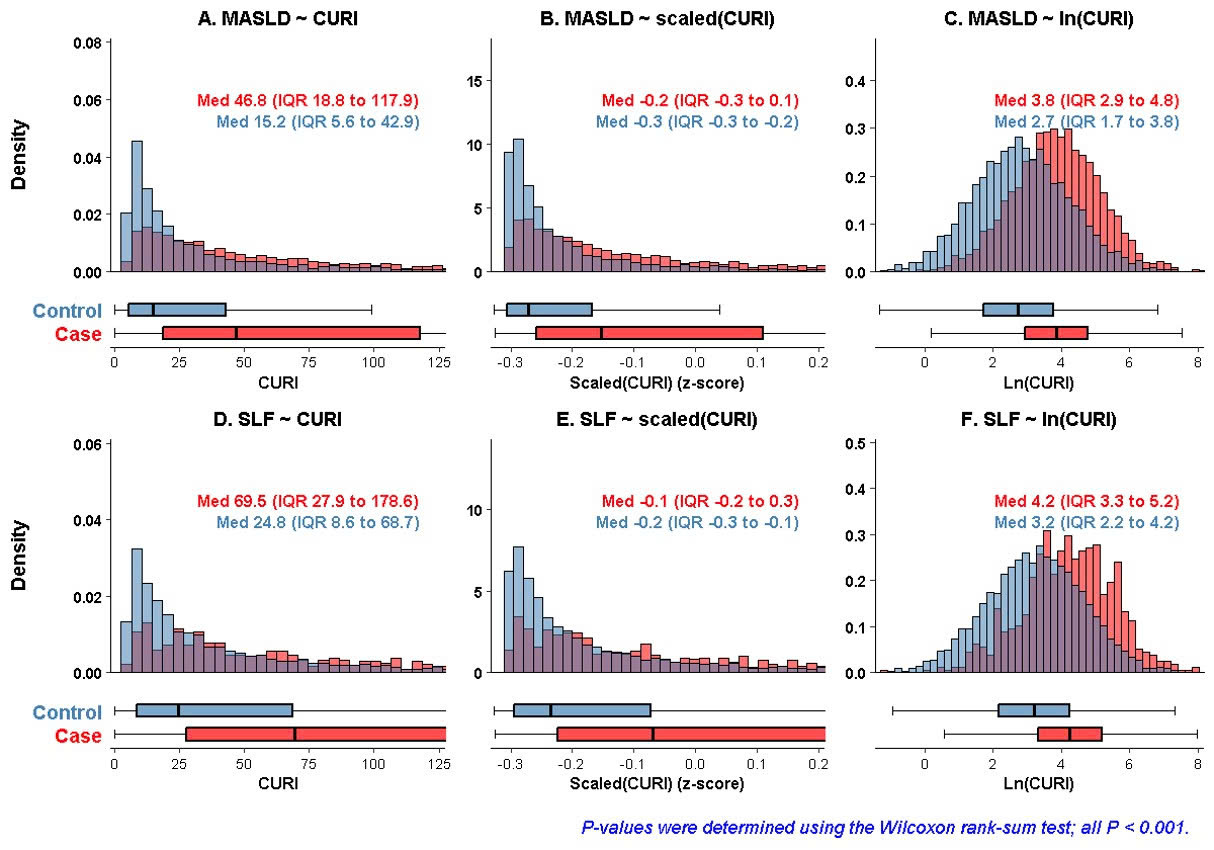

**Supplementary Figure 2. Histogram and boxplot of the CURI with MASLD and SLF.** The cases are participants with MASLD, whereas the controls are non-MASLD participants. Similarly, the cases and controls correspond to participants with and without SLF, respectively.

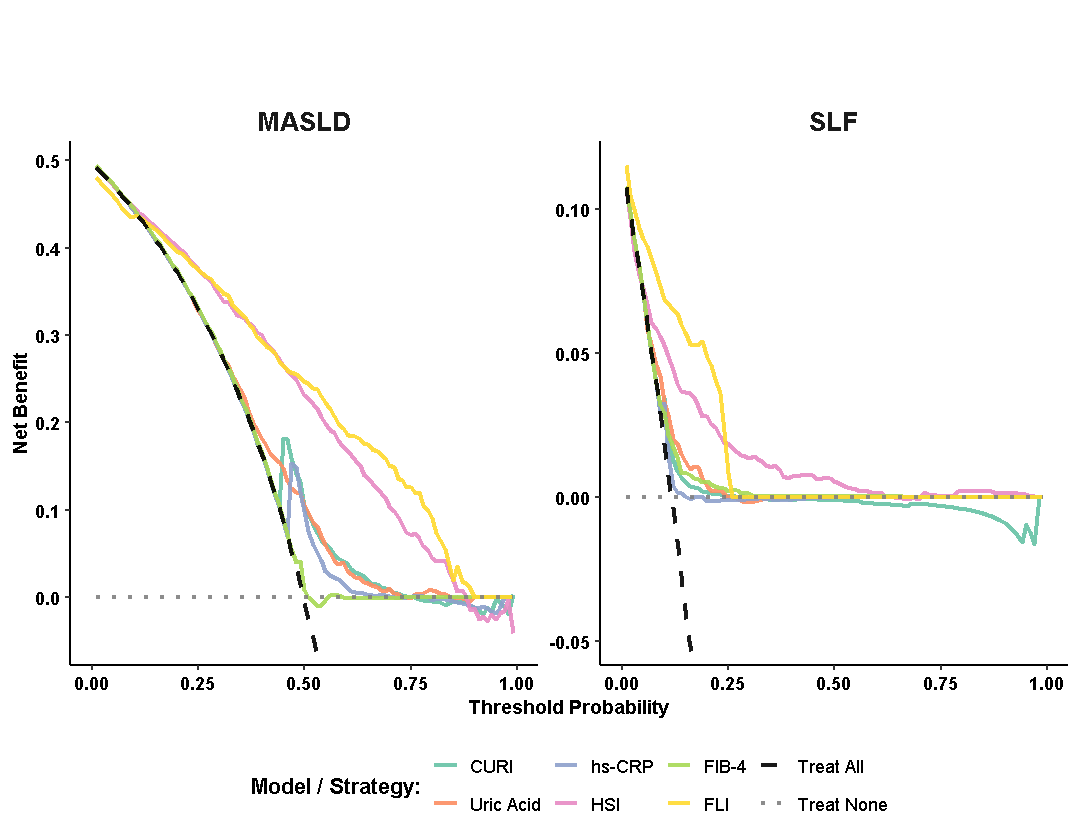

**Supplementary Figure 2. Decision curve analysis of the predictive models for MASLD and SLF. *Abbreviations****: CURI (C-reactive protein to Uric acid ratio index): calculated as hsCRP (mg/dL) × [uric acid (mg/dL)]³; HSI (Hepatic Steatosis Index): 8 × (ALT/AST ratio) + BMI (+2, if female; +2, if type 2 diabetes); FLI (Fatty Liver Index): (e^(0.953 × ln(triglycerides) + 0.139 × BMI + 0.718 × ln(GGT) + 0.053 × waist circumference - 15.745))/(1 + e^(0.953 × ln(triglycerides) + 0.139 × BMI + 0.718 × ln(GGT) + 0.053 × waist circumference - 15.745)) × 100; FIB-4 (Fibrosis-4 Index): [Age (years) × AST (U/L)]/[Platelets (10⁹/L) × √ALT (U/L)]; CI: Represents the value of hsCRP (high-sensitivity C-reactive protein), measured in mg/dL; URI: Represents the value of uric acid, measured in mg/dL.*

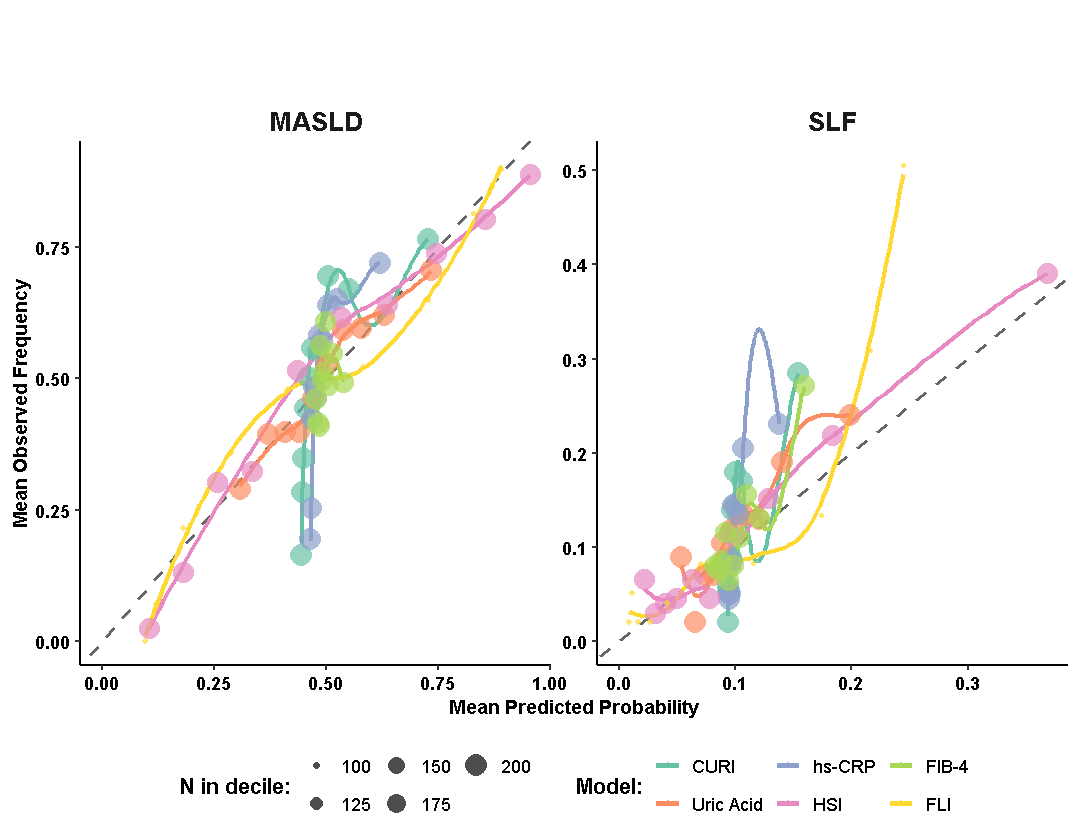

**Supplementary Figure 3. Calibration plots of the predictive models for MASLD and SLF. *Abbreviations****: CURI (C-reactive protein to Uric acid ratio index): calculated as hsCRP (mg/dL) × [uric acid (mg/dL)]³; HSI (Hepatic Steatosis Index): 8 × (ALT/AST ratio) + BMI (+2, if female; +2, if type 2 diabetes); FLI (Fatty Liver Index): (e^(0.953 × ln(triglycerides) + 0.139 × BMI + 0.718 × ln(GGT) + 0.053 × waist circumference - 15.745))/(1 + e^(0.953 × ln(triglycerides) + 0.139 × BMI + 0.718 × ln(GGT) + 0.053 × waist circumference - 15.745)) × 100; FIB-4 (Fibrosis-4 Index): [Age (years) × AST (U/L)]/[Platelets (10⁹/L) × √ALT (U/L)]; CI: Represents the value of hsCRP (high-sensitivity C-reactive protein), measured in mg/dL; URI: Represents the value of uric acid, measured in mg/dL.*
